# What to do when everything happens at once: Analytic approaches to estimate the health effects of co-occurring social policies

**DOI:** 10.1101/2020.10.05.20205963

**Authors:** Ellicott C. Matthay, Laura M. Gottlieb, David Rehkopf, May Lynn Tan, David Vlahov, M. Maria Glymour

**Affiliations:** Center for Health and Community, University of California, San Francisco, San Francisco, California (Ellicott C. Matthay, Laura M. Gottlieb, May Lynn Tan, M. Maria Glymour); Department of Epidemiology and Population Health, Stanford School of Medicine, Palo Alto, California (David Rehkopf); and Yale University School of Nursing, New Haven, Connecticut (David Vlahov)

## Abstract

Social policies have great potential to improve population health and reduce health disparities. Thus, increasing empirical research seeks to quantify the health effects of social policies by exploiting variation in the timing of policy changes across places. Multiple social policies are often adopted simultaneously or in close succession in the same locations, creating co-occurrence which must be handled analytically for valid inferences. Although this is a substantial methodological challenge for studies aiming to isolate social policy effects, limited prior work has systematically considered analytic solutions within a causal framework or assessed whether these solutions are being adopted. We designated seven analytic solutions to policy co-occurrence, including efforts to disentangle individual policy effects and efforts to estimate the combined effects of co-occurring policies. We leveraged an existing systematic review of social policies and health to evaluate how often policy co-occurrence is identified as a threat to validity and how often each analytic solution is applied in practice. Of the 55 studies, only 17 (31%) reported checking for any co-occurring policies, although 36 (67%) used at least one approach that helps address policy co-occurrence. The most common approaches were: adjusting for measures of co-occurring policies; defining the outcome on subpopulations likely to be affected by the policy of interest (but not other co-occurring policies); and selecting a less-correlated measure of policy exposure. As health research increasingly focuses on policy changes, we must systematically assess policy co-occurrence and apply analytic solutions to strengthen future studies on the health effects of social policies.

## INTRODUCTION

Social policies are promising mechanisms to improve population health and reduce health disparities. Analyses of the health effects of social policies routinely leverage policy changes occurring in one or multiple different places at different times, with differences-in-differences or similar study designs (1). In epidemiology and related fields, empirical health research using these methods has proliferated rapidly and yielded important findings (2–4). However, the validity of this approach is threatened when multiple related policies are adopted simultaneously or in close succession in the same jurisdiction. Bundles of related policies with similar potential health effects are often changed together, creating “co-occurrence” that must be addressed analytically for valid inference. Specifically, analyses that do not account for co-occurring policies are likely to be confounded, while analyses that incorporate measures of co-occurring policies can encounter imprecise or unstable estimates and bias resulting from data sparsity (5, 6).

While a rich literature exists on confounding and consequent data sparsity generally (7–12), several aspects of the policy co-occurrence problem make it important to consider separately from issues that arise with other exposures. By nature, policymaking may create correlations among policy variables that are much stronger than those typically observed in non-policy studies (13–15). Governments may respond to the desires of their constituents by adopting multiple related policies at the same time. For example, a state that moves to overhaul its social safety net is likely to change multiple related policies (e.g. income support and food insecurity benefit generosity) at the same time. The most promising analytic solutions may also be different. For example, if a set of policies are always adopted together, then estimating their combined effect is informative for real-world decision-making, whereas analyses of the combined effect of an exposure of interest and closely related confounders may be less useful. Additionally, some data sparsity problems can be addressed by increasing sample sizes, but policy studies are typically based on a small, fixed set of jurisdictions. Meanwhile, stronger theories or substantive knowledge about how a specific social policy functions could guide causal analyses evaluating the health effects of mediating variables (16). For example, if it is understood that compulsory schooling laws affect all-cause mortality by increasing educational attainment for some individuals, then such laws could serve as an instrument for studying the effects of changing educational attainment on health (17). Thus, the policy co-occurrence problem presents distinct challenges and possible analytic solutions beyond typical confounding.

In many policy domains, adopting groups of policies as a set is common (see Part 1 in this paper series). In these cases, researchers can implement a variety of study designs or statistical strategies to address potential bias or imprecision resulting from policy co-occurrence. Among these approaches, an overarching distinction is whether the approach aims to disentangle the effects of individual policies, or conceptualizes the co-occurring policies as a group and evaluates their joint effects. For a given study, either approach may be policy-relevant, depending on whether the goal is to deliver actionable evidence on the effects of a single policy or on the effects of a set of policies that would likely be adopted together.

To date, most approaches to handling social policy co-occurrence have been ad hoc. Applied studies in fields including epidemiology, political science, and health economics have acknowledged the issue by critiquing existing studies that have failed to account for co-occurring policies or by employing specific analytic solutions (18–23). Methodological work in specific fields such as environmental epidemiology, statistics, substance use, political science, and economics have also discussed individual analytic solutions relevant to these domains (6,8,11,24–28). The problem of multicollinearity is widely recognized in econometrics as a threat to causal inference (29–32), but to our knowledge, none have specifically addressed applications to research on the health effects of social policies. In this paper, we aimed to address the need for a systematic assessment of the analytic solutions that are applicable to research on the health effects of social policies, how often these solutions are used in practice, and the tradeoffs to consider in selecting an approach.

This is the second paper in a series on the social policy co-occurrence problem. The first paper demonstrated that co-occurring social policies are pervasive and that adequate adjustment for co-occurring policies is likely to substantially reduce the precision of estimated effects. Given this, delineating methods appropriate for this context is a high priority for the next generation of research on the health effects of social policies. Here we aim to describe analytic strategies prior researchers have adopted to address social policy co-occurrence with the goal of attaining valid inferences. We categorized these approaches based on the type of causal question they answer (e.g. the effect of an individual policy on a population subgroup versus the effect of a bundle of policies on the overall population). Using the sample of social policy evaluations developed in Part 1 of this series, we measured the proportion of studies in which authors assessed policy co-occurrence and the proportion applying each of the different analytic approaches to account for policy co-occurrence. We discuss the advantages and disadvantages of each approach and provide guidance on selecting among them.

## METHODS

### Identification of social policy studies

We leveraged an existing sample of studies on the health effects of social policies to review common strategies for addressing policy co-occurrence. The details of this review are described in Part 1 of this paper series. Briefly, we selected a multidisciplinary set of journals that publish health-related social policy research and are leading journals in their respective fields (*American Journal of Public Health, American Journal of Epidemiology, Journal of the American Medical Association, New England Journal of Medicine, The Lancet, American Journal of Preventive Medicine, Social Science and Medicine, Health Affairs, Demography,* and *American Economic Review*). We screened all 6,794 articles published in these journals in 2019 and included all original, empirical studies aiming to estimate the causal effects of one or more social policies on health-related outcomes (N=55). “Social policies” were defined as non-medical, population-based or targeted policies adopted at a community or higher level, and hypothesized to affect health or health inequalities via changes in social or behavioral determinants. We defined “health-related outcomes” broadly, to include morbidity, mortality, health conditions, and factors such as smoking, homelessness, and sales of unhealthy products. Given our focus on social interventions, we excluded studies that pertained to health care, health insurance, interventions delivered in the clinical setting, medications, or medical devices, including studies of the Affordable Care Act or Medicaid expansion.

### Categorization of analytic approaches

Our framework (Table 1) applies to research questions about the health effects of one particular policy (the “index” policy) in a defined target population. We assume investigators have identified relevant co-occurring policies that might confound the index policy.

**Table 1:** Types of analytic approaches to address policy co-occurrence with corresponding causal research questions

| # | Overall approach | Analytic approach | Corresponding causal research question |
| --- | --- | --- | --- |
| 1 | Effect of individual policy of interest | Adjust for co-occurring policies | What is the effect of the policy of interest on the health outcome? |
| 2 |  | Restrict the study sample to the region of common support | What is the effect of the policy of interest on the health outcome in the restricted sample? |
| 3 |  | Define the outcome on subpopulations likely to be affected by the index policy but not other co-occurring policies | What is the effect of the policy of interest on the health outcome in the subpopulation? |
| 4 |  | Select a less-correlated measure of policy exposure | Example: How does a more generous version of the policy of interest affect the health outcome, compared to a less generous version of the policy interest? |
| 5 |  | Use formal Bayesian methods | What is the best estimate of the effect of the policy of interest on the health outcome, considering both prior knowledge on policy effects and the observed data on policies and outcomes? |
| 6 | Combined effects of multiple policies | Identify and evaluate the impacts of policy clusters | Example: What is the effect of adopting all policies in the cluster versus no policies in the cluster on the health outcome? |
| 7 |  | Use an overall policy stringency or generosity score | What is the effect of differing levels of overall policy stringency or generosity on the health outcome? |

A priori, we designated seven categories of analytic approaches that researchers could adopt in the face of policy co-occurrence, based on the causal question each method could answer (Table 1). We identified these approaches by reviewing multidisciplinary scientific literature on the study of co-occurring exposures, consulting with experts, and drawing on methods used in our own fields of research. We focused on methods that apply to study designs leveraging policy changes occurring in different places and different times, including aggregate or multi-level differences-in-differences and panel fixed effects. We defined “co-occurring” policies as policies whose adoption or implementation was correlated in space and time with an index policy (i.e. places and times with the index policy are also likely to have the “co-occurring” policy) and that likely affected the health outcome under study. This co-occurrence could be at multiple jurisdictional levels (e.g. cities within states), although single-jurisdiction-level studies were the norm.

### Approach 1: Adjust for co-occurring policies

If co-occurrence of related policies with the index policy is not severe (see Part 1 of this paper series), the researcher can adjust for measures of the other policies—for example, by controlling for co-occurring policy measures in a regression. Under conventional assumptions, the resulting estimand corresponds to the effect of the index policy on the health outcome. This approach will often rely on some degree of model-based extrapolation, because not all possible combinations of policies actually occur. It is incumbent on the investigator to confirm that any extrapolation is well-founded in theory or evidence.

As an example, Raifman and colleagues estimated the effect of state same-sex marriage laws on adolescent suicide attempts using a differences-in-differences analysis while controlling for policies banning sexual orientation-based employment discrimination (33). Because co-occurrence between the two types of policies was only moderate, regression adjustment for the co-occurring was sufficient to isolate the index policy.

Policy co-occurrence will be severe if a co-occurring policy aligns perfectly or nearly perfectly in place and time with the policy of interest. After adjusting for co-occurring policies, there will be insufficient independent variation in the index policy left to study, giving extremely imprecise estimates. The only analytic solutions are based on modifying the research question. Approaches 2-7 involve such alternative research questions and corresponding analytic approaches to assessing the impact of the index policy.

### Approach 2: Restrict the study sample to the region of common support

The issue of policy co-occurrence can be conceptualized as a form of strong confounding of the index policy by the other co-occurring policies. This confounding and consequent data sparsity result in a lack of common support in the data, also known as a violation of the positivity assumption (7). Positivity violations occur when some confounder strata do not have variation in the exposure— for example, because the confounding policy and index policy are always adopted as a set. This situation can be resolved by restricting the analysis to the confounder strata for which there is variation in the index policy, i.e., the data region of “common support”. Similar to Approach #1, this approach usually involves adjusting for co-occurring policies, but here, extrapolation is avoided by restricting the study sample. This approach changes the target population, so the corresponding causal question refers to the effect of the index policy on the health outcome in the restricted sample, and results are only generalizable to the population represented by the restricted sample. In the extreme, if an index policy and co-occurring policy are always adopted as set, then there may be no region of common support and alternative approaches must be considered (e.g. evaluating the combined impacts of a bundle of policies).

One way to implement this approach is to restrict the study sample to a subpopulation for whom exposure to the non-index co-occurring policies does not vary. If policies that might confound the association of interest do not vary within a particular study population, then they cannot cause confounding. For example, many households are eligible for multiple social welfare programs including the Supplemental Nutrition Assistance Program (SNAP) and the Special Supplemental Nutrition Program for Women, Infants, and Children (WIC). This makes it difficult to disentangle program effects. Liu and colleagues addressed this co-occurrence by studying outcomes for dual beneficiaries of SNAP and WIC versus WIC alone (34). Those without WIC could be considered “off-support” and are excluded, so all study participants are WIC beneficiaries. This approach changes the target population, so the corresponding causal question refers to the effect of the index policy (e.g. SNAP) on the health outcome in the restricted sample (e.g. WIC beneficiaries) and results generalize to the population represented by the restricted sample.

When there are many confounder strata, one accessible way to assess positivity and identify the region of common support is using propensity scores. In the context of assessing policy impacts, the propensity score (35) is the probability of adopting the index policy, given the confounding policies. Units that are “on-support” are those with propensity scores within the range of observed propensity scores both for units adopting the index policy and for units not adopting the index policy. A wide variety of matching and weighting methods involve using propensity scores to identify and restrict to the region of common support (10, 36). Numerous variations on this restriction have also been proposed, including restricting to units with propensity scores within a prespecified range (e.g. 0.1 to 0.9) or dynamic optimization procedures for selecting propensity score cutoffs (7–9).

An alternative approach to using propensity scores is to directly restrict the sample based on the distribution of the co-occurring policies themselves. Several approaches have been proposed, including restricting to units inside the convex hull of the covariate space defined by the secondary policies (11, 37), restricting to a sufficiently data-dense, rectangular region of the covariate space defined by the co-occurring policies (38–40), or tree-based methods (see for example (41)). These approaches are less common, but most can be readily implemented using existing software. In all cases, assessing the region of common support helps ensure that estimates are not relying on extrapolation to policy combinations which are never observed. The restricted study population should be well-defined, so that the investigator can transparently describe the places and times to which the results apply (38).

Chang and colleagues applied this approach to study the impacts of prescription drug monitoring programs (PDMPs) and pill mill laws using a comparative interrupted time series analysis (42). Rather than using all states in the analysis, some of which implemented other opioid policy changes in concert with PDMPs or pill mill laws, the authors restricted their analysis to Florida, which adopted PDMP and pill mill laws, and Georgia, which had a similar policy profile to Florida but did not implement the index policies during the study period. The authors determined that the combination of PDMPs and pill mill laws were associated with reductions in high-risk opioid prescribing for the Florida population.

### Approach 3: Define the outcome on subpopulations likely to be affected by the index policy but not other co-occurring policies

Identifying health effects that are specific to the index policy can be achieved by changing the outcome measure to one that is both closely aligned with the index policy and unlikely to be affected by other co-occurring policies. In particular, if the outcome is focused on a particular population subgroup (e.g. defined by age, gender, place, or time) that is likely to be most-affected by the index policy and unlikely to be affected by other co-occurring policy changes, study results can provide pointers to the impacts of the individual policy.

For example, changes in state Earned Income Tax Credit (EITC) policies have often co-occurred with other changes in other social welfare policies such as SNAP (43). Rehkopf and colleagues took advantage of the fact that EITC cash benefits are typically delivered in February, March, and April, while other benefits do not have the same seasonal dispersal pattern, to examine the association of EITC policies with health (44). They used a differences-in-differences approach to compared health outcomes that can change on a monthly basis (e.g. health behaviors, cardiovascular and metabolic biomarkers) for EITC-eligible versus non-eligible individuals in months of income supplementation versus non-supplementation. The authors were thus able to measure some potential short-term health impacts of EITC independent of other social welfare policies that do not have this seasonality.

This approach can be further strengthened by incorporating falsification tests or negative control analyses. Rehkopf and colleagues strengthened their findings by confirming that treating non-EITC transfer months as “treated” months produced null findings, and by confirming null associations for outcomes that do not change on a monthly basis.

### Approach 4: Select a less-correlated measure of policy exposure

Studies that use binary (0 or 1) characterizations of policy adoption are widespread in studies of the health impacts of social policies. However, more detailed characterizations of individual policies—for example, the amount of funding allocated, benefit generosity, participation rate, or population reach of a program; the size of a tax; or the number of years a policy has been in place—can deliver policy measures that are less correlated with other related policies, or opportunities to examine dose-response effects among jurisdictions adopting a policy. For example, adoption of more generous unemployment benefits, in terms of dollar amounts and durations for different types of households, tend to change in tandem with other worker protection and leave policies, but researchers have effectively disentangled effects of unemployment benefits by leveraging continuous measures of maximum allowable unemployment benefit levels across states (45, 46). Similar approaches have been taken to studying the effects of alcohol taxes (47, 48), tobacco taxes (49), and EITC benefit generosity (50, 51).

One useful modification to this approach is to study factors that may specifically mediate the relationship between the index policy and the health outcomes. For example, Matthay and colleagues generated evidence on the impacts of policies regulating gun shows by examining the impacts of gun show events themselves on firearm-related injuries in differing policy environments (52). Similarly, the impacts of medical cannabis provisions allowing supply through dispensaries (as opposed to home-cultivation) can be quantified by studying the direct effects of dispensaries on health (53, 54).

Studying mediators may also offer the opportunity to identify policy effects via the Front Door Criterion (16), a rarely-used alternative to confounder-control or instrument-based methods (4). If all the pathways by which the index policy affects the outcome can be measured, and there are no unmeasured confounders of the index policy-mediator relationship or of mediator-outcome relationship (conditional on the index policy), then the effect of the index policy on the outcome can be identified without measuring the co-occurring policies. For example, Bellemare and colleagues used the Front Door Criterion to estimate the effect of authorizing Uber and Lyft ride sharing with strangers on tipping drivers (55). They propose that they only way in which sharing authorization affects tipping is through whether a ride is actually shared (the mediator). This mediator is used as a tool to estimate the effect of sharing authorization while circumventing confounders of sharing authorization such as rider experience, mood, and social preferences. Although applications of the Front Door Criterion remain rare, a similar approach could be applied to social policy evaluations.

### Approach 5: Use formal Bayesian methods

Bayesian methods can be used to integrate information gleaned from other approaches to addressing policy co-occurrence; Bayesian methods can also be used as a way to treat estimation problems arising from policy co-occurrence without linking to other approaches we have mentioned. Several approaches to addressing policy co-occurrence depend on incorporating prior knowledge about the policies, determinants of the outcome, or hypothesized mechanisms of effect. For example, researchers may apply judgements about which policies affect the outcome or modify other policy processes. These insights can guide decisions about which policies need to be controlled and how. Bayesian methods offer a formal statistical method to incorporate prior knowledge about the plausible effects of the co-occurring and index policies, and to combine these with newly gathered empirical data.

When used alone, Bayesian methods can help address estimation issues and recover precision when highly co-occurring policies lead to convergence problems or imprecision. In particular, Bayesian approaches can stabilize estimates (i.e. address data sparsity-related problems of imprecision and sensitivity to different model specifications and influential data points) by constraining the effect sizes or interaction effects among policies and “shrinking” coefficients towards the specified prior distributions (22). This can be done without changing the set of adjustment variables, without restricting the study sample, and without changing the exposure or outcome measure. This approach is common in the environmental epidemiology literature as way to study multiple co-occurring exposures such as air pollutants (6, 26).

Harper used a Bayesian differences-in-differences approach to estimate the effects of adopting stronger enforcement of state seat belt laws on motor vehicle crash deaths (56). Because other road safety policies may also affect motor vehicle crash deaths and change in tandem with seat belt enforcement policies, Harper also adjusted for laws controlling maximum speed limits, blood alcohol concentration limits, graduated driver’s license programs, and annual state policy per capita as a proxy for traffic safety enforcement. Because these variables are correlated across states and years with each other and with seat belt enforcement policies, adjusting for them in a frequentist analysis reduces the precision of the estimated effect of interest—a major problem resulting from co-occurring policies. Harper enhanced precision by applying a Bayesian approach, drawing on existing evidence of the effects to seat belt laws to place empirical priors on the estimated effect of seat belt enforcement policies.

### Approach 6: Identify and evaluate the impacts of policy clusters

If a set of policies are typically adopted as a group, the effect of the combined set of policies may be the most pertinent parameter to estimate. By conceptualizing policy clusters as the exposure of interest, the investigator can preserve the original target population and outcome measure. For example, if two or more policies are highly co-occurring, it may be possible to estimate their combined impact (e.g., comparing health outcomes if both policies were adopted versus if neither policy were adopted) (57). Policy clusters can be defined based on substantive or policymaking considerations; this is useful if decisionmakers are considering adopting a set of policies. Alternatively, numerous data-driven clustering algorithms are applicable. Clusters or categories are defined based on how frequently policies co-occur (i.e., the extent to which policies co-occur in the same place and time). Methods include hierarchical cluster analysis, latent class analysis (LCA), or principal components analysis (PCA) (6,58,59). Clusters might also be defined based on the strength of the relationship with the outcome—for example using supervised PCA (6, 26). No one algorithm is considered optimal for all settings (6, 26).

Among data-driven algorithms, it is common to distinguish between “variable-centered” methods that group similar policy variables (e.g. PCA) and “person-centered” methods that group similar observations (e.g. LCA). The underlying mechanics of variable-centered and person-centered approaches are distinct, but both ultimately result in a small set of variables that summarize the policies to which each observation is exposed. This smaller set of variables is then used to assess health impacts. Erickson and colleagues used LCA to classify US states based on their position on 18 alcohol control policies (60). The analysis categorized each state into one of four unordered groups which the authors interpreted as: weak except serving policies, average, strong for underage use, and strong policies overall. State policy category was then associated with levels of past-month alcohol consumption.

### Approach 7: Use an overall policy stringency or generosity scor

If the investigator is interested the effects of the overall policy environment on health, one method is to use a summary score of the stringency or generosity of a set of policies. In comparison with Approach 6, this approach similarly involves reducing many policy variables to a few, but it typically focuses on creating an ordering along a pre-defined unidimensional scale such as stringency. In contrast, policy clusters are usually unordered and defined based on the co-variation amongst the policy measures themselves without regard for underlying characterizations such as stringency, although such characterizations may be applied after the fact when interpreting or describing the clusters. This approach also differs from Approach 4—in which one might characterize a single policy on a continuous scale to help disentangle the effects of that policy from other policies— because here we characterize a collection of policies with respect to their likely combined impact.

A simple way to apply this approach is to sum the number of policies in the set that apply in each place and time. Policies must be coded in the same direction so that the presence of more policies indicates greater restrictiveness, or vice versa. This method is easy to operationalize, but it implies that all policies carry equal weight and are interchangeable in achieving health effects.

A more sophisticated application is to weight policies based on existing evidence or expert opinion about the strength of the relationship with the outcome. This may be based on efficacy, restrictiveness, implementation, enforcement, enforceability, reach, or other metrics. Investigators have applied this approach in literature on firearm policy (61, 62), alcohol policy (20,63–65), and cannabis policy (66, 67). Although there are an infinite number of ways a set of policies can be ranked or weighted, use of systematic methods can enhance rigor and replicability. For example, the Delphi technique is a structured communication approach to elicit consensus from a panel of experts and can be used to rank or score policies based on stringency or effectiveness (68, 69). Assigned weights are typically outcome-specific—for example, weighting state alcohol policies with different levels effectiveness for binge drinking versus impaired driving and for adults versus youth (20, 63). Investigators can also explore different methods of weighting in sensitivity analyses (63).

### Data extraction and analysis

For each social policy study, we reviewed the full text. Our main focus was on the primary analytic specification, which we understood to be the authors’ leading approach to estimate the effect of the social policy for the health outcome(s) of interest. We also reviewed any sensitivity analyses reported in the main text. We assessed: (a) the overall analytic approach (e.g. differences-in-differences); (b) whether the authors reported checking for any co-occurring policies related to the health outcome of interest that might pose a threat to validity; (c) the authors determination of whether or not any co-occurring policies did, in fact, threaten validity (e.g. based on their analysis or prior literature); (d) whether there was any other indication that co-occurring policies exist for the study’s application (e.g. a co-occurring policy mentioned in the limitations); (e) if policy co-occurrence was identified as at threat, what analytic strategy the authors used to address it; and (f) any other aspects of the analytic strategy that may help address co-occurring policies, whether they were identified as a threat or not. We also documented whether studies utilized any approaches to address policy co-occurrence not identified a priori. We then tabulated these characteristics.

## RESULTS

We assessed 55 studies of social policies encompassing diverse topics, countries, and jurisdictional levels (70–125) (Appendix Table 1). Studies included, for example, a comparative interrupted time series evaluation of the impacts of lowering the blood alcohol concentration limit for drivers on road traffic accidents in Scotland (73) and a differences-in-differences analysis of the effects of state paid family leave policies on breastfeeding (98). The most common domains were poverty and social welfare policies such as the Supplemental Nutrition Assistance Program (14 studies); food and beverage policies such as sugar-sweetened beverage taxes (6 studies); firearm restrictions (5 studies); unemployment, sick leave, and pension benefit policies (4 studies); tobacco control (4 studies); alcohol control (4 studies); and immigration (4 studies).

Figure 1 presents a flowchart of the included studies, broken down by whether the authors evaluated policy co-occurrence and used techniques to address policy co-occurrence. Of the 55 studies, 4 involved methods for which assessing policy co-occurrence was not relevant: One involved a national policy with randomized rollout across village clusters, and for three others, the primary research question was about the overall policy environment and authors employed policy stringency scores. Of the remaining 51 studies, only 17 reported checking for at least one co-occurring policy. Of these 17, 10 reported identifying at least one co-occurring policy while 7 suggested that no co-occurring policies were a threat. For example, in a study of state texting-while-driving bans and traffic injuries, the authors acknowledged that administrative license suspension, speed limits, seatbelt requirements, and graduated driver licensing laws had also changed over the study period and might affect traffic injuries; they therefore controlled for measures of these policies in their differences-in-differences analysis (112). In contrast, a study of the effects of tuition-free primary education on access to family planning and health decision-making evaluated potentially co-occurring paid family leave policies (86); they determined that these policies did not substantially co-occur with tuition-free primary education but acknowledged that there may be other unmeasured co-occurring policies. Of the 34 studies that did not report checking for at least one co-occurring policy, 5 had some other indication that policy co-occurrence may be a threat.

**Figure 1:**
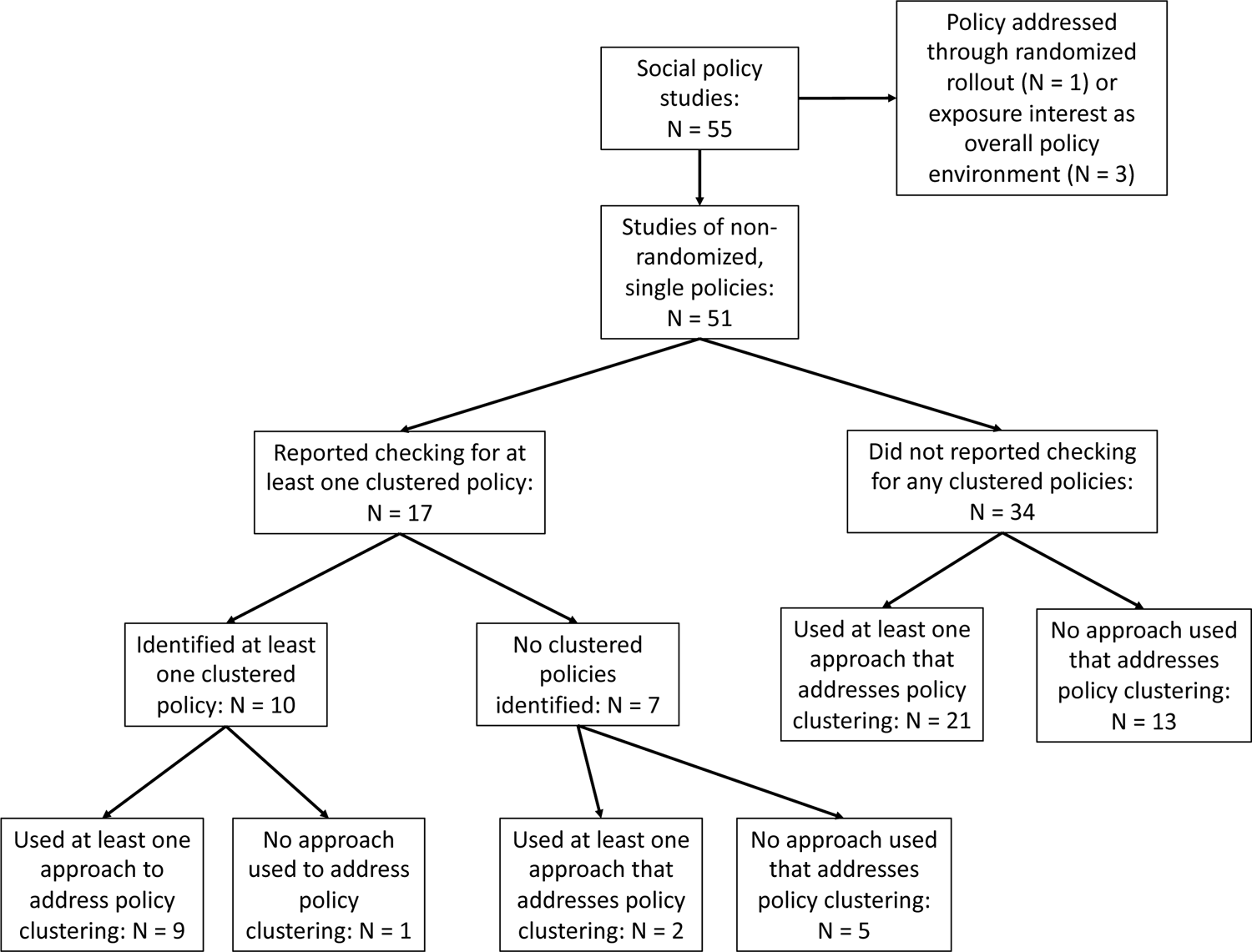
Flowchart of included social policy studies by evaluation of policy co-occurrence and use of techniques to address policy co-occurrence

Table 2 summarizes the analytic methods used in each study, irrespective of whether the authors checked for co-occurring policies. Overall, 36 of the 55 studies (65%) incorporated at least one approach that addressed policy co-occurrence. Among studies that utilized at least one approach, the most common approaches were: adjusting for co-occurring policies (18 studies, 50%); defining the outcome on subpopulations likely to be affected by the index policy (but not other co-occurring policies) (14 studies, 39%); and selecting a less-correlated measure of policy exposure (7 studies, 19%). Twelve (33%) used more than one approach. None used formal Bayesian methods. Two studies reported no co-occurring policies but, without naming co-occurring policies as the motivation, nonetheless applied at least one approach that helps address co-occurring policies (Figure 1).

**Table 2:** Overall analytic methods and approaches used to address policy co-occurrence in social policy studies

| **Approach to address policy co-occurrence** | **Study design** |  |  |  |  |  |  |  |  |  | **Total studies using approach** |
| --- | --- | --- | --- | --- | --- | --- | --- | --- | --- | --- | --- |
|  | DID | Panel FE | CITS | Synth control | Before-after | Regress | PSM | Rand wedge | IV | Sim model |  |
| Adjust for co-occurring policies | 9 | 5 | - | - | 1 | 1 | - | - | 2 | - | 18 |
| Restrict the study sample to the region of common support | - | - | - | 1 | - | 1 | - | - | - | - | 2 |
| Define the outcome on subpopulations likely to be affected by the index policy but not other co-occurring policies | 8 | - | - | 1 | 1 | 3 | 1 | - | - | - | 14 |
| Select a less-correlated measure of policy exposure | 3 | 1 | - | - | - | 1 | - | - | 2 | - | 7 |
| Use Bayesian methods | - | - | - | - | - | - | - | - | - | - | 0 |
| Identify and evaluate the impacts of policy clusters | 3 | - | - | - | - | 0 | - | - | 1 | - | 4 |
| Use an overall policy stringency or generosity score* | 1 | - | - | - | - | 2 | - | - | - | - | 3 |
| No method used | 5 | 2 | 1 | 1 | 7 | 1 | 0 | 1 | - | - | 20 |
| **Total studies using study design** | 12 | 6 | 1 | 2 | 9 | 7 | 1 | 1 | 3 | 1 |  |

Legend: Cells indicate the number of studies using the designated study design and approach to address policy co-occurrence. “-” indicates that no studies applied the given combination of study design and approach to address policy co-occurrence. Margins do not add up to the total number of studies included in the review because some studies used multiple approaches to address policy co-occurrence. Abbreviations: DID: differences-in-differences. Panel FE: Panel fixed effects. CITS: Comparative interrupted time series. Synth. control: Synthetic control. Regress: Other regression without place-specific controls. This study design refers to analyses that estimated the effect of the policy by modeling the outcome as function of some policy exposure measure, but without employing one of the other structured designs such as differences-in-differences or panel fixed effects. PSM: Propensity score matching. Rand. wedge: Randomized stepped wedge. IV: Instrumental variables. Sim. model: Simulation model. \* Studies that used an overall policy stringency or generosity score were those in which the primary research question was about the overall policy environment.

Studies employed a range of designs, the most common being differences-in-differences (12 studies), before-after (e.g. t-tests or interrupted time series; 9 studies), and other regression approaches without place-specific controls (hereafter, “other regression approaches”; e.g. multilevel regression of an overall policy stringency score on an individual-level health outcome without fixed effects; 7 studies) (Table 2). Studies using differences-in-differences, panel fixed effects, or other regression approaches were more likely to use at least one approach to address policy co-occurrence than studies using before-after designs.

Of the 10 studies that explicitly reported identifying one or more co-occurring policies, 9 attempted to address it (Figure 1). Of the 41 studies that did not check for or identify policy co-occurrence, 23 nonetheless used at least one technique that helps address policy co-occurrence. Several studies used multiple approaches in the same analysis. For example, a study of the effects of losing SNAP benefits controlled for participation in WIC and Temporary Assistance for Needy Families (TANF) program (Approach 1); excluded those with potential concurrent benefit changes in Supplemental Security Income (SSI) (Approach 2); and compared outcomes among SNAP participants who lost benefits to similar individuals with continuous benefits (Approach 3) (95).

## DISCUSSION

Co-occurring policies are common and can threaten the validity of studies of the health effects of social policies. In this paper, we describe seven analytic approaches to address policy co-occurrence. The approaches either seek to disentangle the effects of individual policies or estimate the combined effects of clusters of policies. Using a sample of contemporary studies on the health effects of social policies, we found that potential policy co-occurrence is frequently unidentified and unaddressed: only 33% of studies reported checking for policy co-occurrence as a potential threat to validity and only 65% incorporated any approach that helps to address policy co-occurrence, regardless of whether the authors checked for policy co-occurrence. Several studies that estimated the individual effect of the index policy used multiple, possibly complementary approaches to address co-occurrence in the same analysis; this may further enhance validity, although it is not guaranteed.

In future applied studies, systematically evaluating and reporting on policy co-occurrence would facilitate the evaluation of validity and interpretation of findings. Many studies (35%) did not report any approach to address policy co-occurrence. Authors may not have considered it or may have checked for it but not reported their assessment, particularly if it was not found to be a concern. However, because failure to address policy co-occurrence (if it exists) poses a major threat to validity, readers need to understand if the authors believe that no policy co-occurrence exists or if they believe it has been addressed (and if so, through what analytic strategies). Stringent word limits on many medical and public health journals preclude presenting full analytic exploration of issues such as policy co-occurrence. An openness to incorporating such discussions, at least in appendices, would enhance the rigor and interpretability of social policy studies.

We assessed whether the studies in our sample checked for *any* co-occurring policies, but ideally, researchers would evaluate *all* policies and related social, economic, and political phenomena that co-occur with the index policy and that could affect the health outcome of interest. This is a formidable task, especially given that innumerable policies are continuously being passed at all levels of government, that databases measuring relevant policies often do not exist, or that the policies that may affect an outcome are not fully understood. Policy libraries such as the University of Kentucky Center for Poverty Research state welfare database (43), the Policy Surveillance Program at Temple University (126), and the University of Iowa State Policy Innovation and Diffusion Database (127) are increasingly valuable resources, but they are burdensome to develop and maintain and require infrastructure support. Substantial, regular support for policy surveillance as well as scientific endeavors to link and harmonize large administrative datasets would support these efforts (128–130). Given that diverse policies across numerous disciplines are likely to affect health, interdisciplinary collaboration is also essential to these efforts.

### Tradeoffs between different approaches

This study describes how approaches to address policy co-occurrence have been used practice; a logical next question is which approaches are best-suited for different circumstances. The preferred approach to address policy co-occurrence should be driven by the target causal question (131). Investigators should select the approach that best-answers their causal question, while achieving sufficient accuracy and precision (e.g. based on likely sources of bias and evidence on the precision of different estimators). If an unbiased, precise estimate can be derived with simple adjustment for measures of the co-occurring policies (Approach 1), this option will often be preferable, because it does not change the target causal question or study population. However, if the target causal question is deemed unanswerable due to severe policy co-occurrence, a different analytic approach, potentially corresponding with a modified causal question, is necessary (131). More severe policy co-occurrence may necessitate larger departures from the original causal question. The extent to which alternative approaches change the causal question depends on the application—for example, whether restricting the study sample to the region of common support involves dropping many units or only a few—but Appendix Figure 1 shows an approximate ordering.

Table 3 summarizes tradeoffs of different approaches to addressing policy co-occurrence. Overall, approaches that preserve estimates of the independent effect of the index policy may be particularly useful for decisionmakers comparing specific policy options. However, these approaches generally sacrifice some aspect of generalizability by restricting the analysis to certain populations, subgroups, outcomes, or time periods for which policy effects can be estimated. Results may therefore serve as markers of policy impacts rather than measures of overall impact. Estimating the combined effects of a group of co-occurring policies sacrifices estimates of the independent effects of the index policy, but preserves generalizability to the original target population, outcomes, and time period under study. The preferred approach depends both on what options are viable (i.e. unconfounded, sufficient precision) and which causal question is of greatest interest. For example, if certain combinations of policies are always adopted together, then their independent effects may be neither estimable nor of interest.

**Table 3:** Advantages and disadvantages of alternative approaches used to address policy co-occurrence in studies of the health effects of social policies

| Approach | Advantages | Disadvantages |
| --- | --- | --- |
| *Approaches involving disentangling the effects of individual policies* | Results are informative for decisionmakers interested in whether or not to adopt the index policy of interest. | Most approaches require sacrificing some aspect of generalizability by restricting the analysis to certain populations, subgroups, outcomes, or time periods for which policy effects can be estimated. |
| 1. Adjust for co-occurring policies | Does not requiring changing the original research question. | Only works if policy co-occurrence is not severe (no perfectly aligned policies; sufficient statistical power and independent variation in index policy of interest after controlling for co-occurring policies). |
| 2. Restrict the study sample to the region of common support | Need to be able to identify the region of common support; propensity scores are most common but must be correctly estimated. Supported by a large literature on using propensity scores for analyzing policy effects. Helps ensure that estimates do not rely on extrapolation to policy combinations which are never observed. | Reduces sample size; can harm statistical power; restricts the population to whom the results generalize. If using propensity scores, they must be correctly estimated. |
| 3. Define the outcome on subpopulations likely to be affected by the index policy but not other co-occurring policies | Can isolate individual policy effects in the face of severe policy co-occurrence. Encourages drilling down on the times, places, and people that are most-affected or of greatest interest. | Policy-specific outcomes must exist, be correctly identified (based on existing evidence or theory), and be relevant to the research question of interest. Can inhibit direct comparison of effect estimates from policy alternatives using uniform methods and measures of association. Assumes no spillover effects of the index policy on any comparison or control groups deemed “unaffected” by the index policy. |
| 4. Select a less-correlated measure of policy exposure | Can isolate individual policy effects in the face of severe policy co-occurrence. Encourages drilling down on the hypothesized mechanisms and policy aspects that are most-affected or of greatest interest. | Policy-specific exposures must exist, be correctly identified (based on existing evidence or theory), and be relevant to the research question of interest. Can inhibit direct comparison of effect estimates from policy alternatives using uniform methods and measures of association. |
| 5. Use formal Bayesian methods | Can solve estimation problems without sacrificing the ability to study individual policy effects in the original target population | Does not solve fundamental lack of support in the data. May still rely on extrapolation. Often computationally intensive. Methods and format of |
|  |  | results are less familiar to some audiences. |
| *Approaches involving estimating the combined effects of clusters of policies* | Preserves generalizability of the original target population, outcomes, and time period under study. May answer the most policy-relevant question if certain bundles of policies are always adopted together. | Does not produce estimates of individual policy effects; cannot distinguish which policies in a cluster are driving health effects. |
| 6. Identify and evaluate the impacts of policy clusters | Can provide useful estimates of the combined impacts of realistic policy combinations. | No consensus on optimal methods to identify policy clusters or optimal criteria for selecting a final set of clusters (particularly concerning if effect estimates are sensitive to the choice of clustering) (59). Results can be challenging to interpret when the summary policy measures are weighted combinations of policy variables, as in PCA or factor analysis, or if the clustering algorithm produces many distinct clusters that are difficult to define or interpret. |
| 7. Use an overall policy stringency or generosity score | Summarizes the effect of the overall policy environment. May be the only viable option in the face of severe policy co-occurrence. | Developing weighting schemes can be time-consuming and subjective. Results can be sensitive to the choice of score, score weighting, or score components, unless using data-driven weighting schemes based on the strength of the relationship with the outcome. Implies that two policies are interchangeable in their effects if adopting one or the other results in the same numeric change in the score (possibly unrealistic). |

Among the individual approaches, key considerations include the circumstances in which the approach is feasible (e.g. controlling for co-occurring policies is not possible if policy co-occurrence is severe), the availability of evidence to support making analytic decisions (e.g. on how to use propensity scores, select weighting schemes for policy scores, or choose a clustering method), the extent to which the approach provides evidence that is relevant to the original causal question, ease of implementation, available data and measures, and interpretability of the results (see Table 3 for details). All of the approaches discussed here can also be used to evaluate whether policy co-occurrence is a concern by comparing results of analyses that do not account for policy co-occurrence to results from analyses that do. While none of these approaches will answer identical research questions, findings should generally align and comparison across methods can serve as a robustness check.

For all of the approaches, we note two important limitations. First, none of the seven approaches discussed here are guaranteed to resolve the analytic challenges presented by co-occurring policies. For example, one approach might isolate the effects of the primary policy of interest from some co-occurring policies but not others; another approach might help reduce problems of statistical power arising from policy co-occurrence for one outcome of interest but not another. Second, all of the approaches rely on accurate measurement of all of the relevant policies. Missing or mis-measured policies may lead to bias. Careful attention to the structure and potential impact of measurement error, along with analytic tools such as quantitative bias analysis, can enhance validity (132).

### Limitations

The seven approaches presented here are not an exhaustive list of all analytic solutions that could be applied to address policy co-occurrence and many sub-options exist. However, we did not encounter any other method that addresses policy co-occurrence in our sample of studies. Additionally, this study is based on a systematically gathered set of exemplar studies of the health effects of social policies; a comprehensive review of all studies of the health effects of social policies would be valuable in future research—for example to characterize patterns of methods utilization across journals and disciplines, and to assess whether studies are trending towards more rigorous approaches over time. Finally, as with all studies, there may be some misclassification. In particular, if an analytic approach was applied but not identified as for the purpose of addressing co-occurring policies (or an analogous problem under any other name), we may have missed it.

## Conclusions

Policy co-occurrence plagues most research on the health effects of social policies. In combination with Part 1 of this paper series—which illustrated how to assess the pervasiveness and consequences of policy co-occurrence—this review offers guidance on how to address this challenge. While randomization of policy rollouts can best estimate the causal effects of social policies, when not available, other methods can nonetheless indicate causality. These other methods demand careful selection of the research question and analytic approach and, guided by deep substantive knowledge and creativity, can help to overcome policy co-occurrence and deliver stronger evidence on the health effects of social policies.

## Data Availability

All data used in this study arise from published studies that are publicly available.

## Appendix

**Appendix Table 1:**
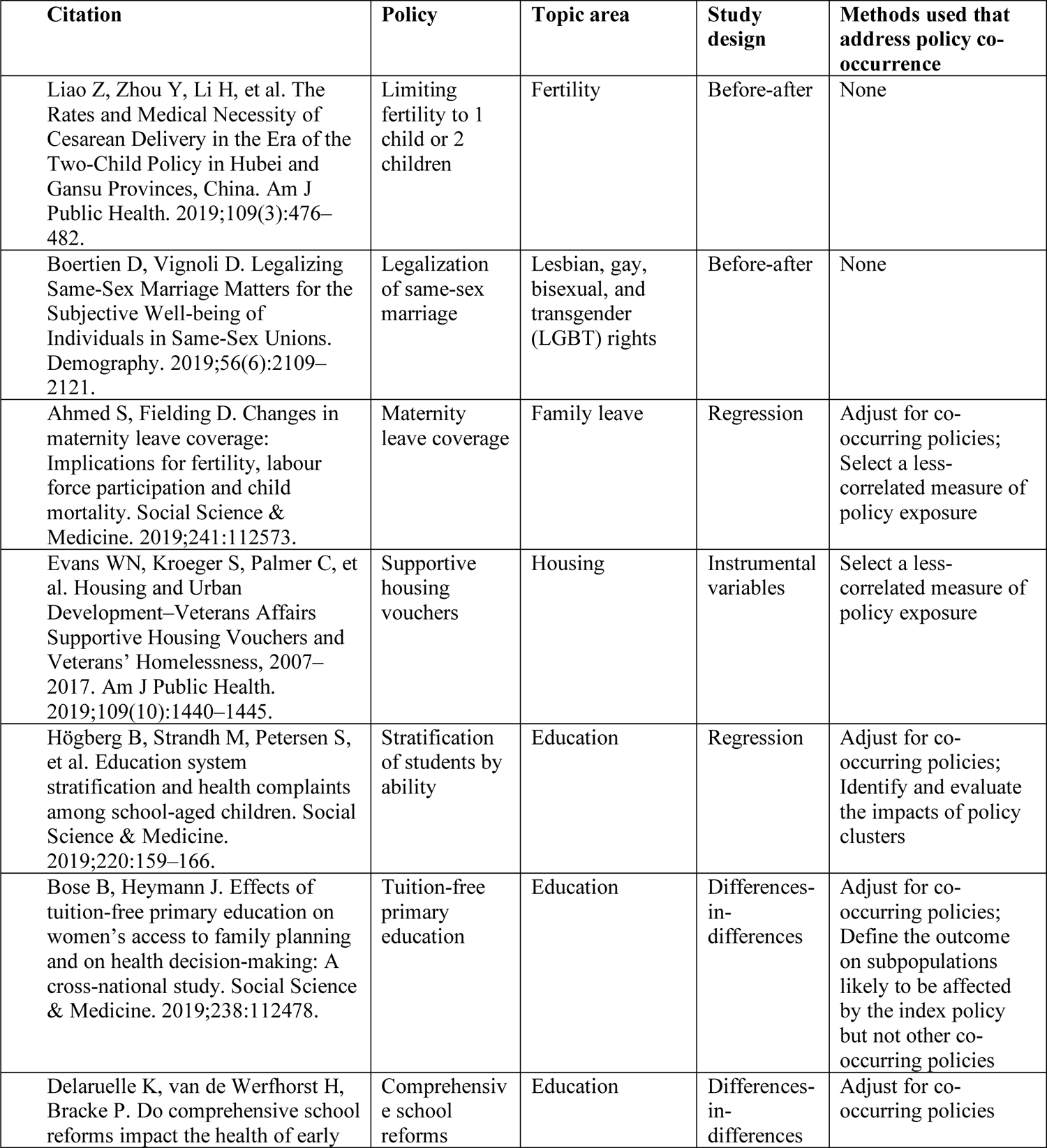
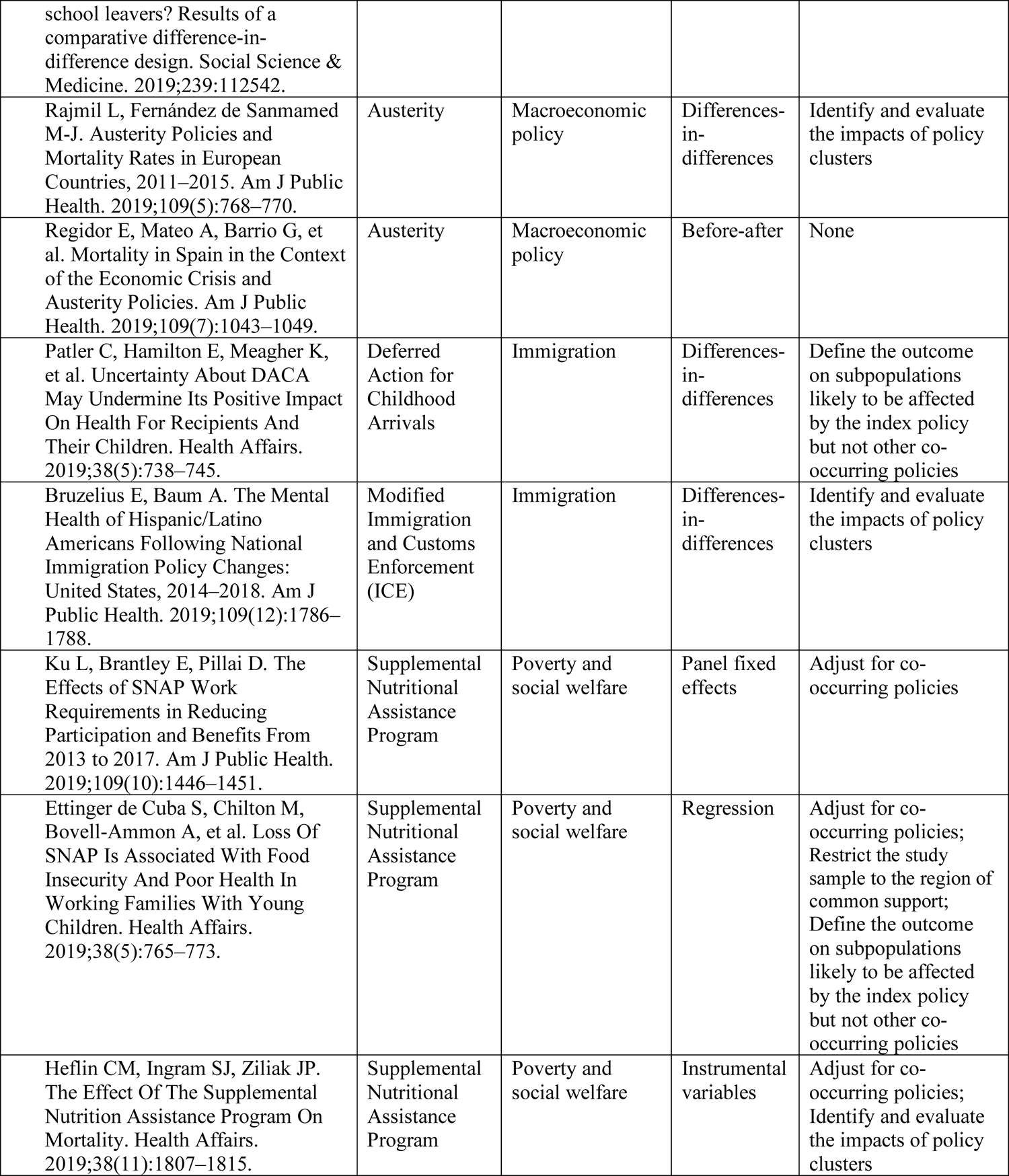
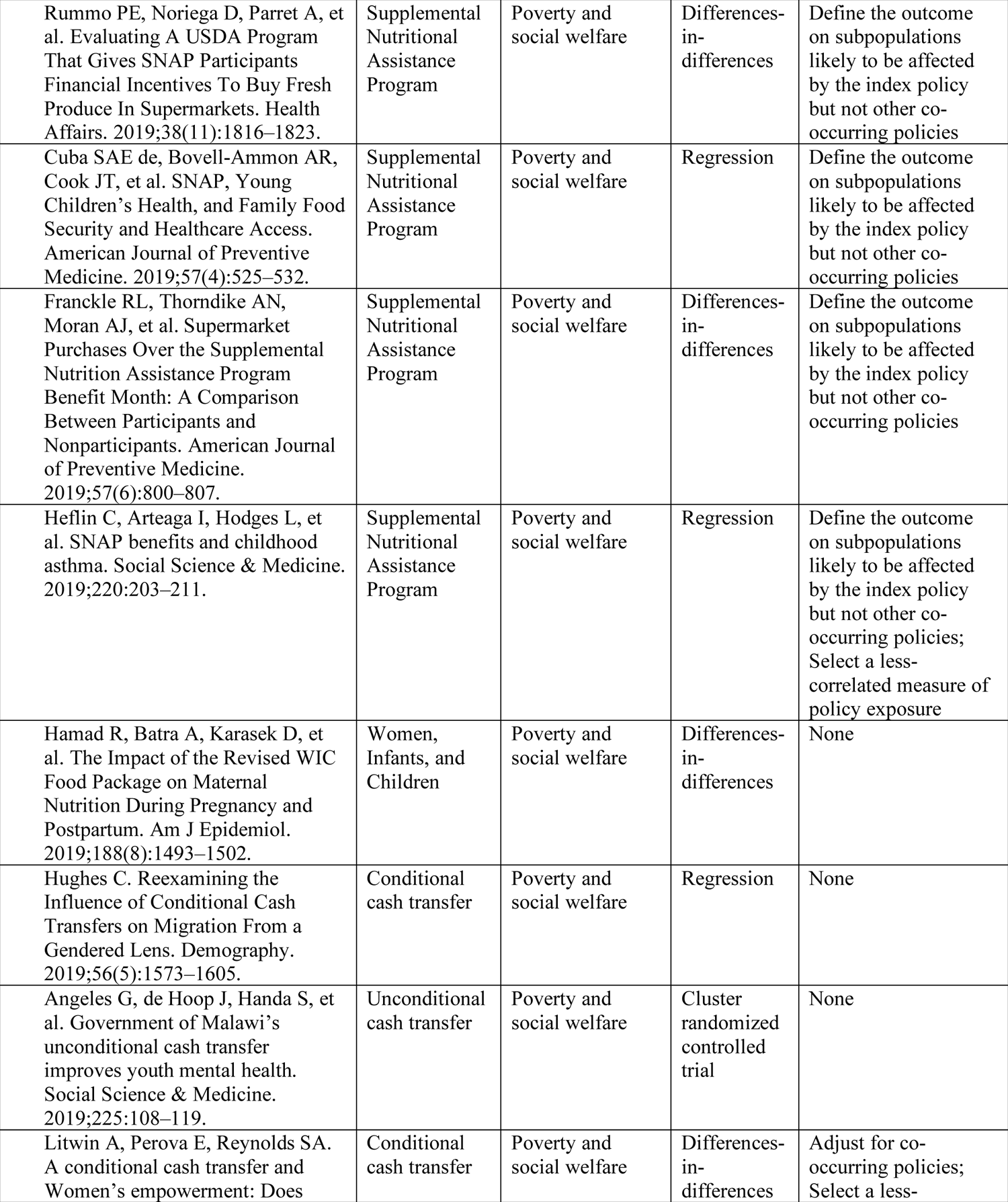
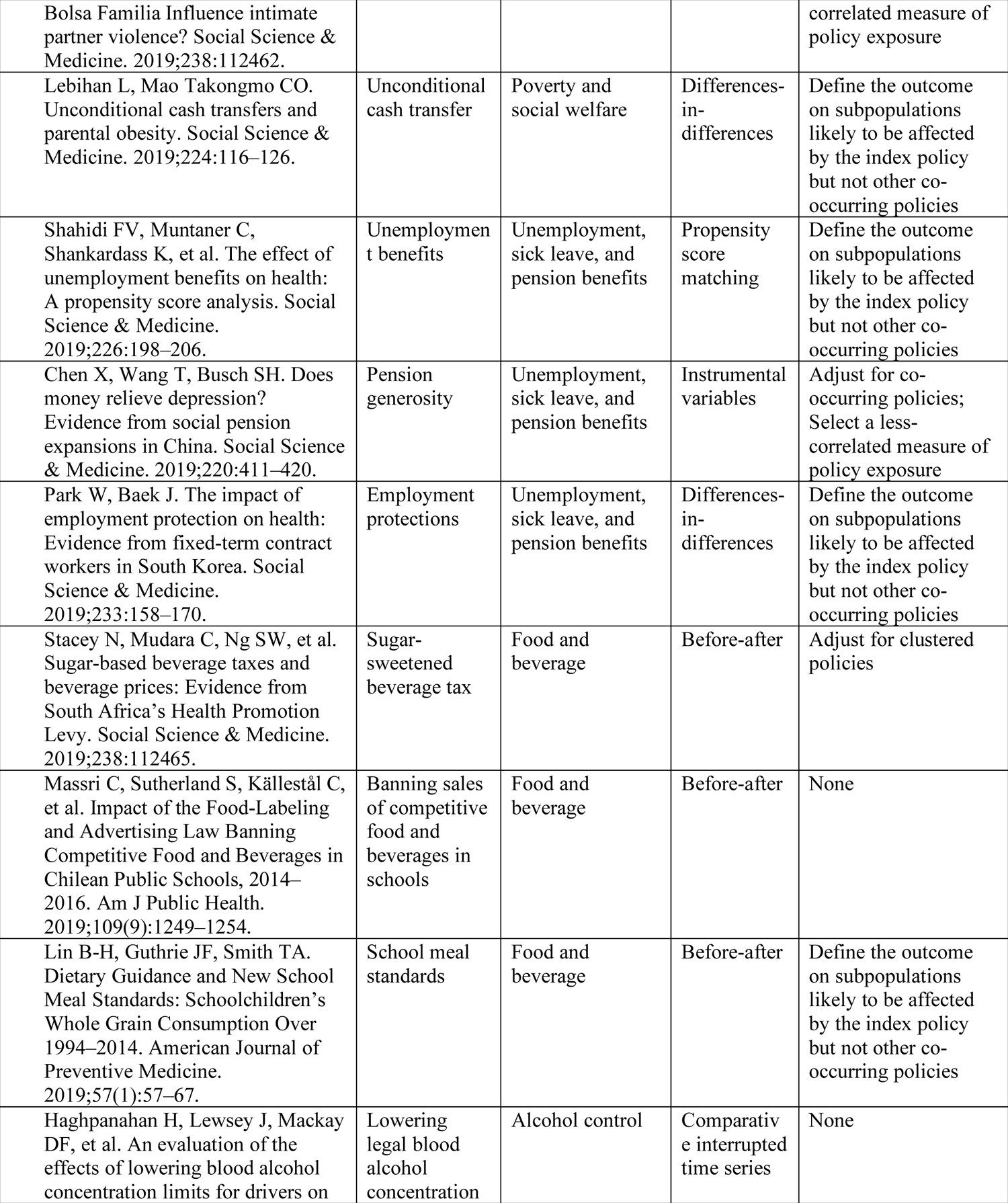
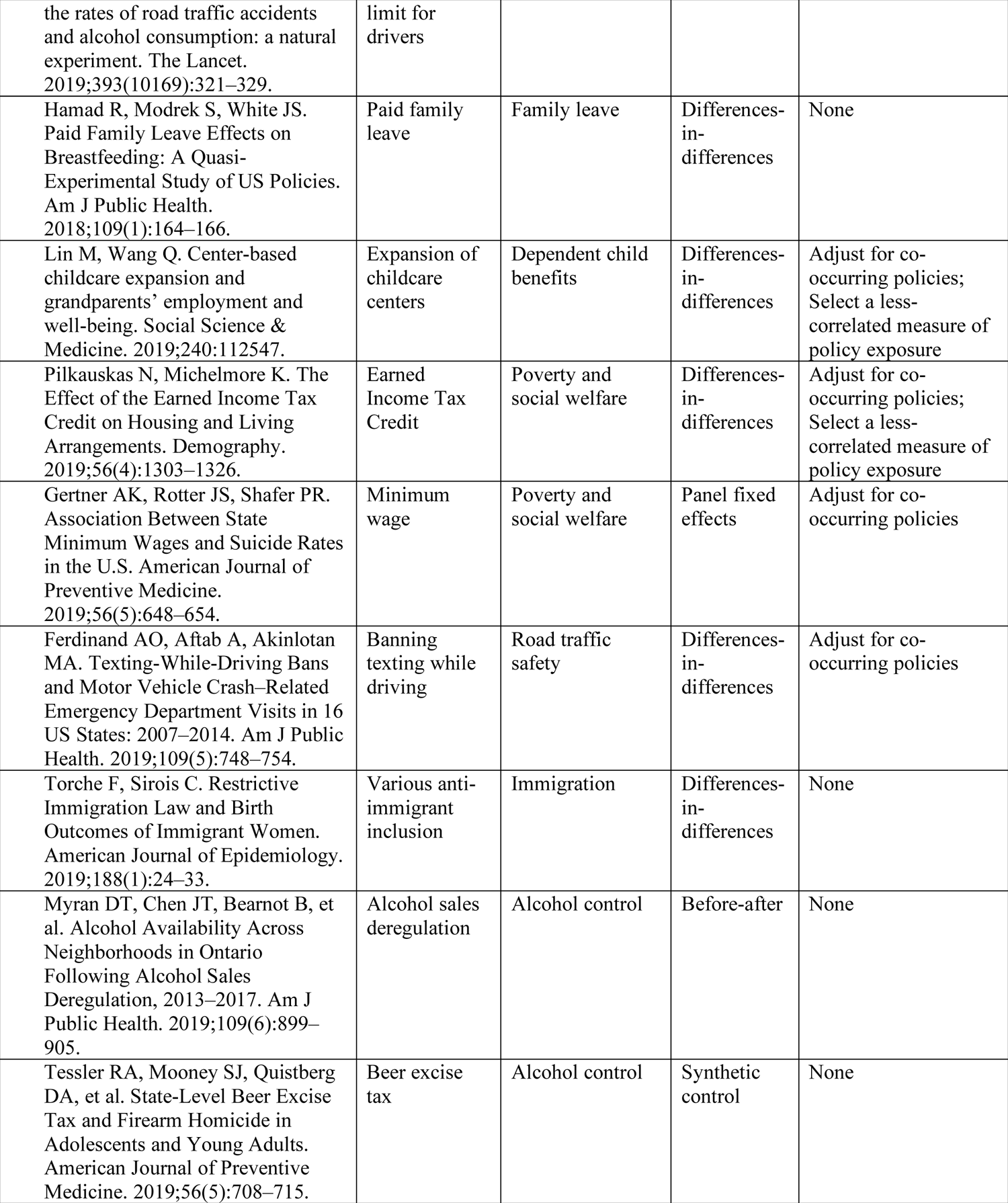
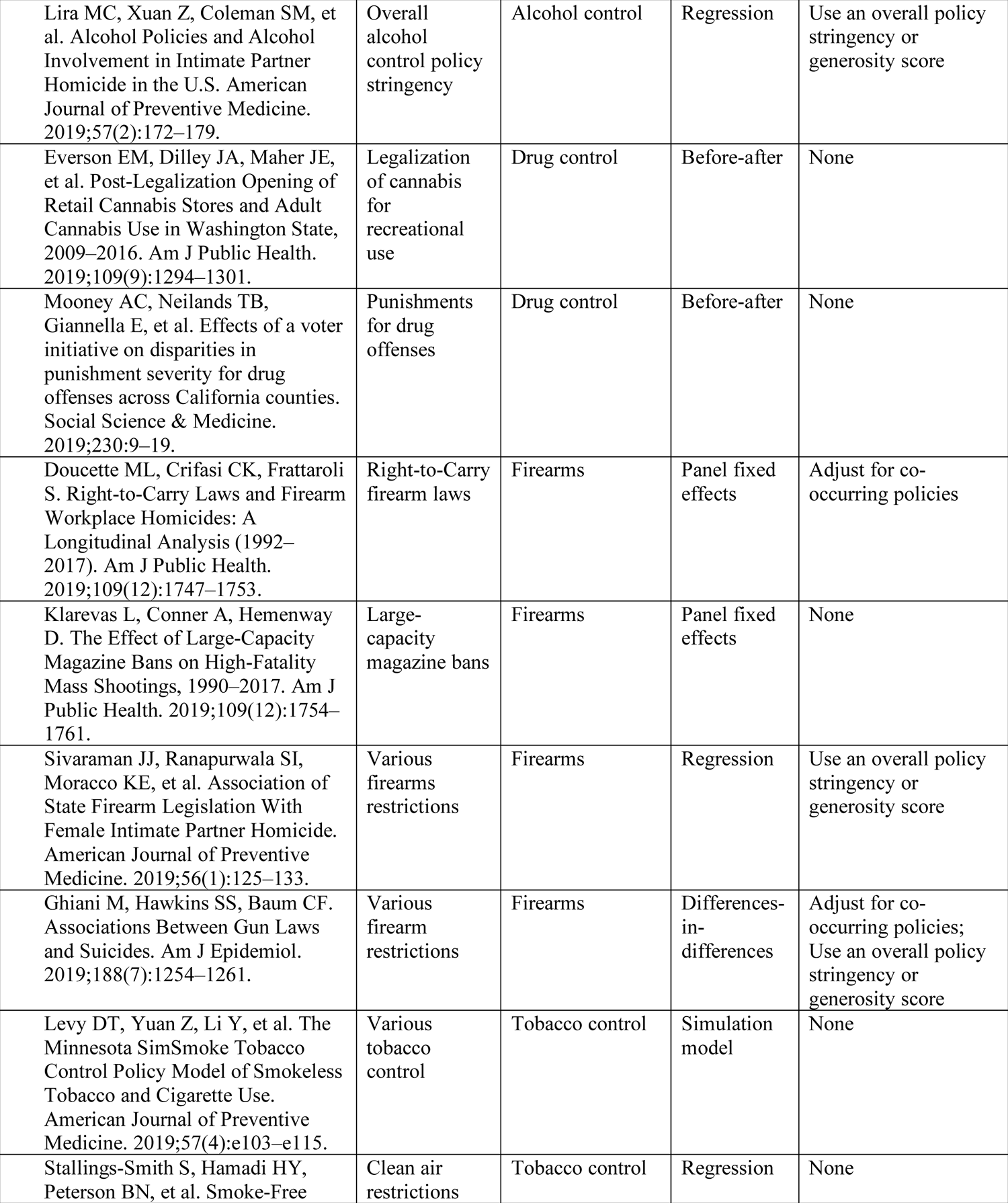
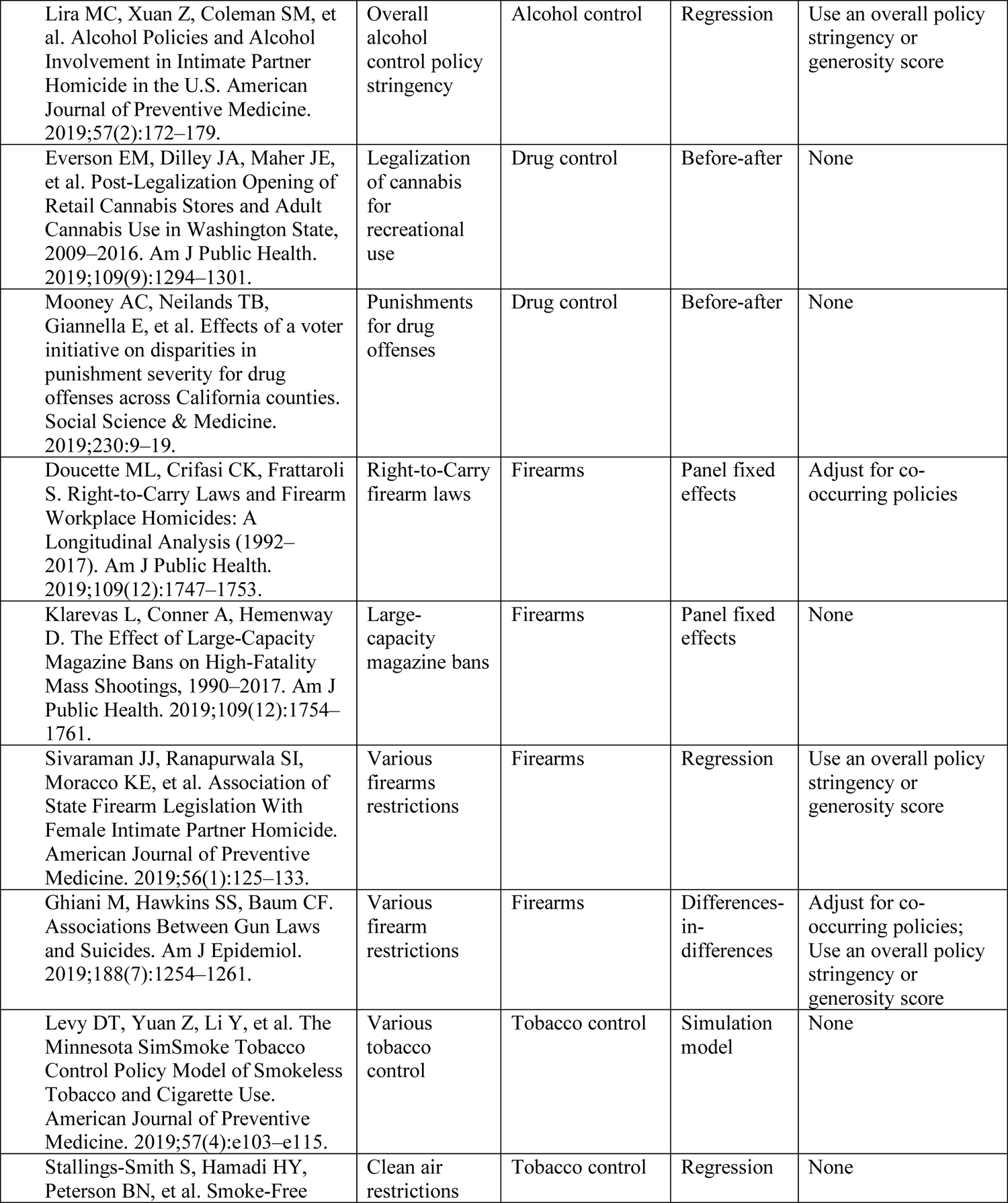
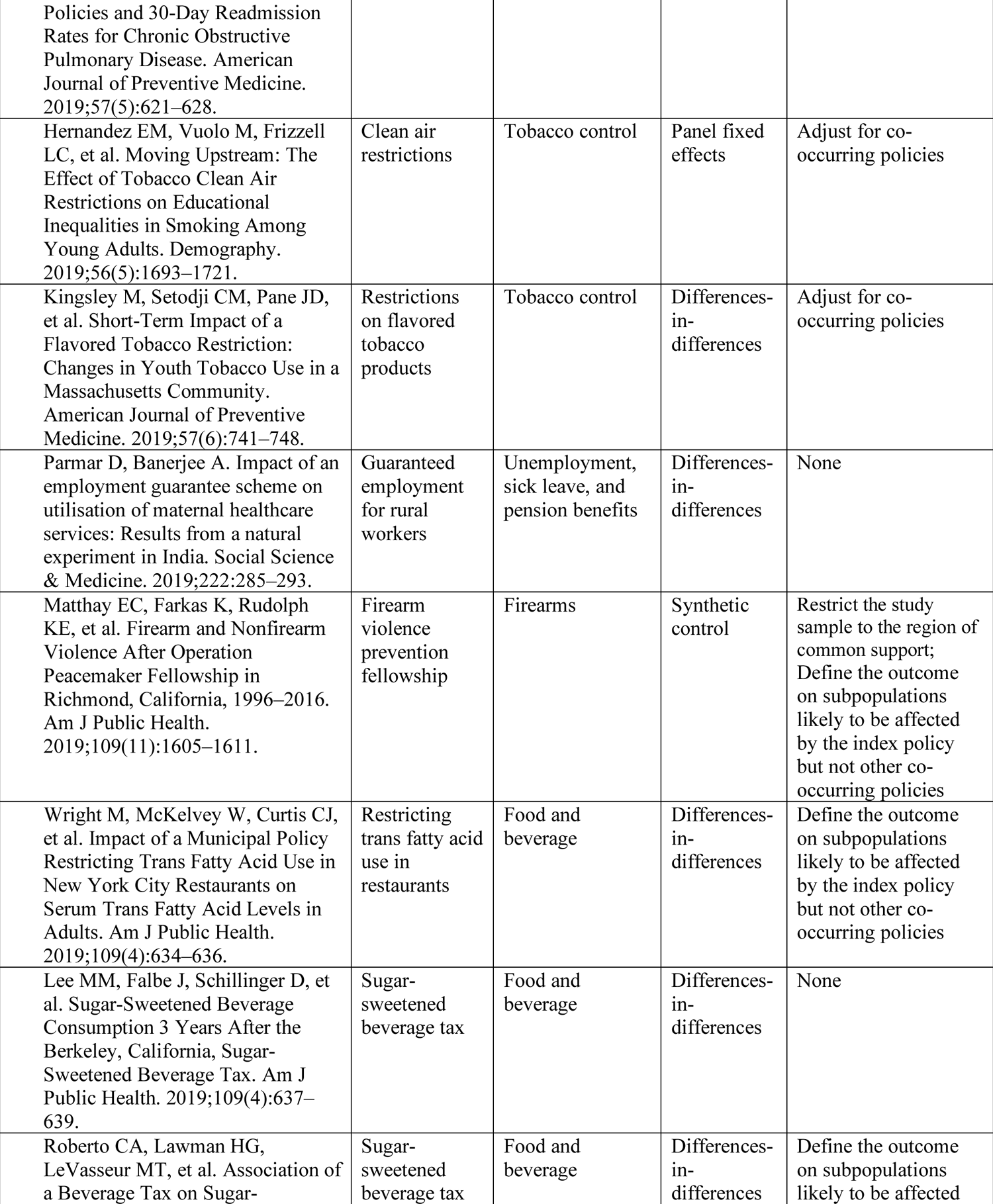
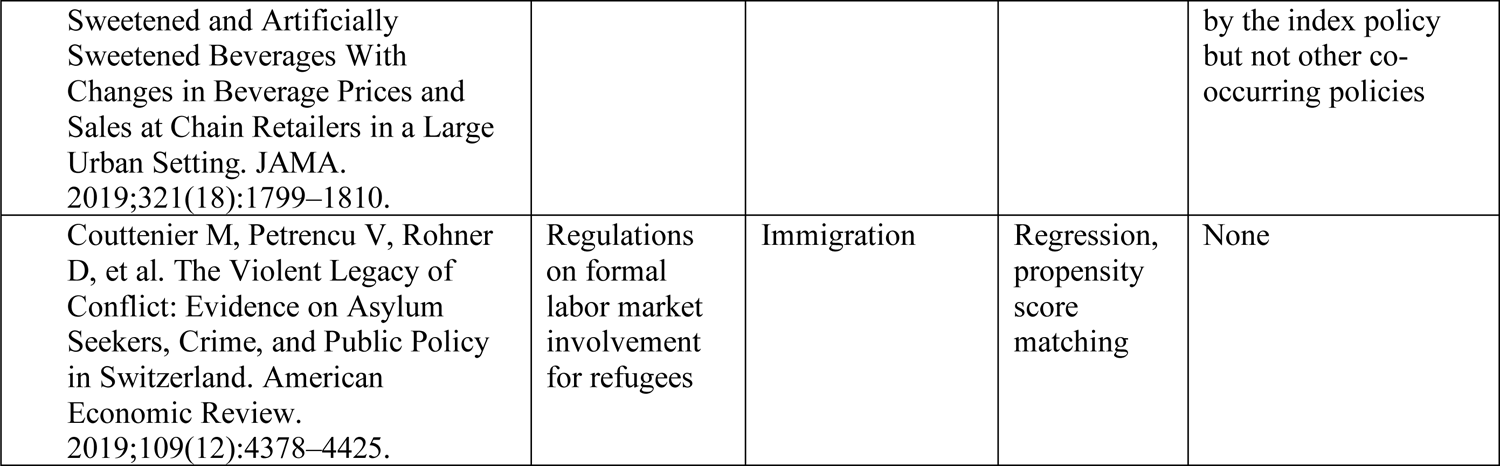
Social policy studies identified in systematic sample

**Appendix Figure 1:**
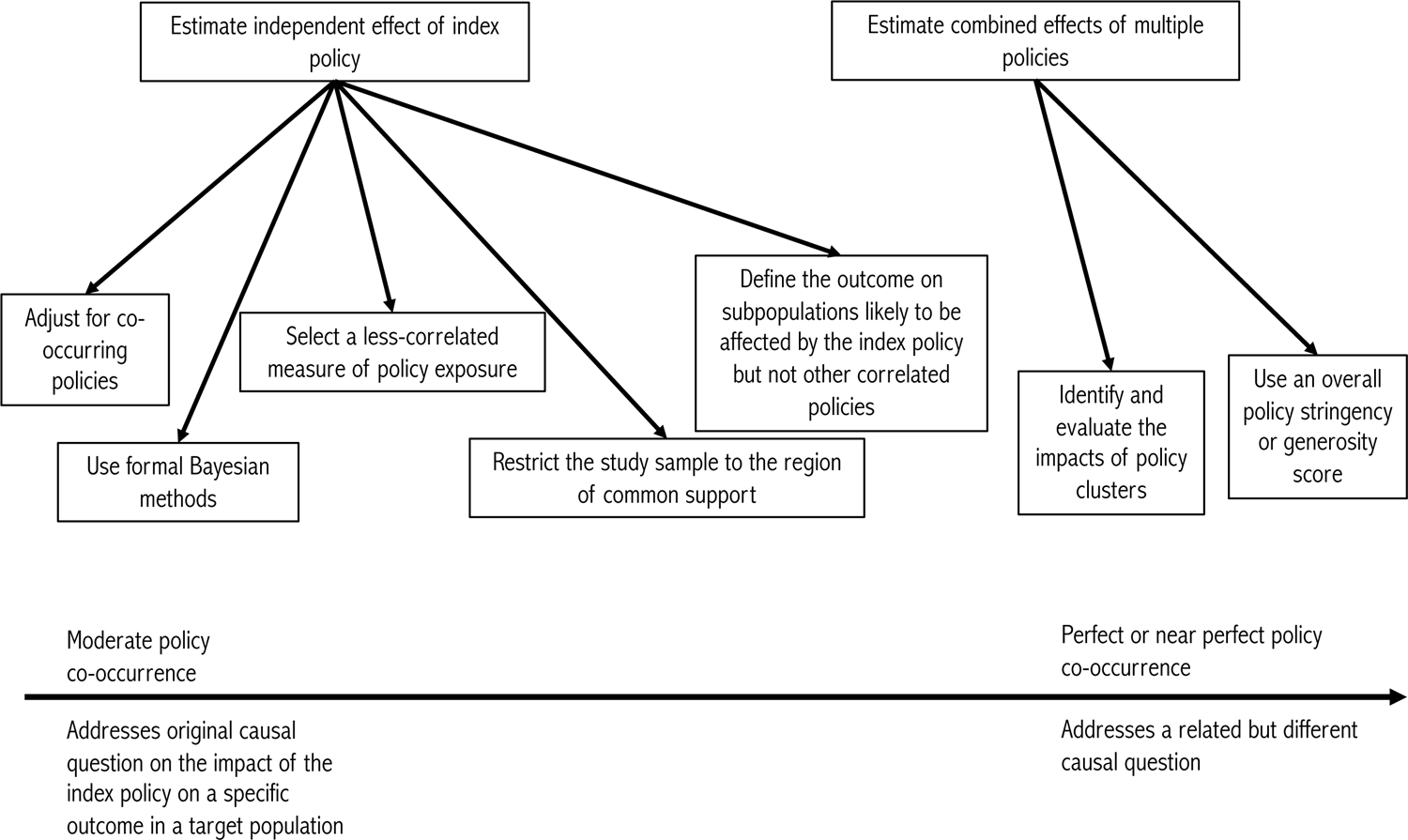
Schema of alternative approaches to address policy co-occurrence by severity of policy co-occurrence and degree of departure from original causal question

